# Counterfactual Analysis of Executable Clinical Decision Logic

**DOI:** 10.64898/2026.08.05.26359737

**Authors:** Camelia Maleki, Yannis Bertrand, Frederik Gailly

## Abstract

Clinical recommendations are often expressed in narrative form, which limits their direct execution, auditability, and patient-specific interpretation. This paper presents a hybrid decision-support framework that combines Decision Model and Notation (DMN), survey-weighted rule-ensemble learning, and counterfactual sensitivity analysis. The framework is evaluated using an NHANES-derived fasting cohort for classification of documented diabetes status. The full fasting analysis cohort contained 2,582 participants, and a non-diagnostic laboratory subgroup, Gate0, contained 2,111 participants. On untouched test data, the rule-ensemble model achieved ROC–AUC and PR– AUC values of 0.959 and 0.873 in the full fasting cohort and 0.861 and 0.499 in Gate0. Four clinically interpretable candidate rules were selected using validation data only. A nonnegative survey-weighted logistic model removed one redundant rule and converted the remaining three binary activations into an auditable DMN score and model-estimated probability. The final DMN achieved ROC–AUC 0.769, PR–AUC 0.153, and Brier score 0.029 in the untouched Gate0 test set. In small rule-defined test subgroups, hypothetical five-unit BMI reductions lowered mean model-estimated probability by 2.40 to 5.89 percentage points when one or more BMI thresholds were crossed. These findings characterize policy sensitivity rather than causal effects and require external validation.

## I. Introduction

Clinical practice guidelines aim to standardize care by translating evidence and expert consensus into decision logic. Their implementation can nevertheless vary across settings and patient profiles, particularly when measurements are incomplete, borderline, or changing. In such situations, clinicians may need not only a recommendation but also an explanation of which feasible changes would alter the output of the decision process [1], [2].

Decision Model and Notation (DMN) provides a structured representation of decision logic that is human-readable, executable, traceable, and auditable. DMN decision tables make conditions, rule-firing paths, and outputs explicit, thereby supporting clinical review and integration with health-data workflows [3], [4]. However, a DMN table normally evaluates the current input state; it does not automatically show how its output would change under feasible modifications.

This study combines three components. First, observational data are used to identify outcome-relevant variables and candidate rule patterns. Second, clinically interpretable candidate rules are represented in an executable DMN collect/sum decision table. Third, selected modifiable inputs are perturbed and the DMN is re-executed to produce transparent “what-if” explanations. Because these changes operate through explicit threshold conditions, their effects can be traced to rule activation and deactivation rather than attributed to an opaque prediction process [5], [6].

The contribution is a validation-separated framework for data-supported DMN construction and counterfactual policy sensitivity. The case study uses an NHANES-derived fasting cohort, applies fasting-subsample survey weights, separates model development from final testing, and reports both the predictive rule-ensemble model and the simpler DMN policy [7]–[9].

## II. Background

Formal representations of clinical knowledge can reduce ambiguity and support consistent execution across information systems [10]. DMN is well suited to this purpose because its input columns describe patient attributes, its rows define explicit conditions, and its output columns specify the resulting decision (Fig. 1). Hit policies determine how multiple matching rules are handled; under a collect/sum policy, the numeric outputs of all matching rules are added [3], [4].

**Fig. 1.**
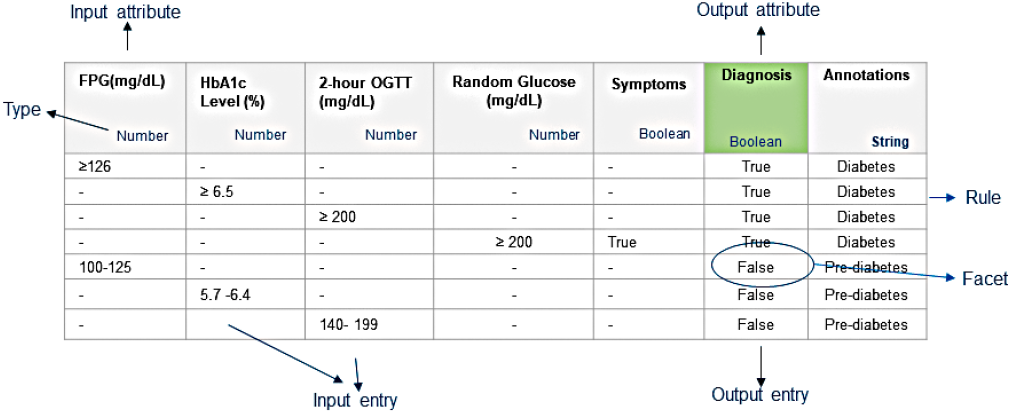
Core elements of a DMN decision table.

Observational data can support clinical decision logic by identifying variables and combinations associated with an outcome [11]–[13]. Such patterns should not be treated as clinical recommendations without expert review; rather, they provide candidate operational rules that can be assessed for plausibility, stability, and alignment with domain knowledge.

Counterfactual explanations describe input changes associated with a different model or policy output [5]. In the present framework, counterfactuals are implemented by changing one modifiable input and re-executing the DMN. This produces a discrete policy-sensitivity analysis: the output changes only when the modified input crosses a threshold used by an active rule. The resulting difference is not a causal treatment effect [14], [15].

## III. Method

Figure 2 summarizes the workflow. Let **x** denote routinely collected clinical and demographic variables, and let *y* ∈ {0, 1} denote the binary diabetes reference label. The framework returns binary DMN rule activations, an additive score, and a model-estimated probability 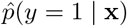.

**Fig. 2.**
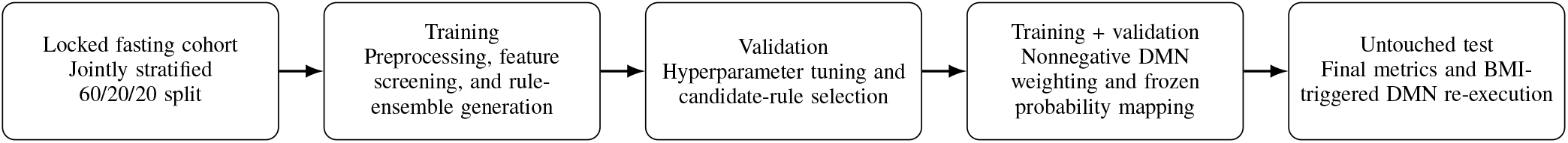
Validation-separated workflow. Training data support feature screening and rule generation; validation data support tuning and candidate-rule selection; the combined development cohort estimates the frozen DMN weights; and the untouched test set is used only for final evaluation and counterfactual policy-sensitivity analysis.

The reference outcome was defined independently of the laboratory thresholds used in the DMN. A positive label was assigned when `DIQ010, DIQ050`, or `DIQ070` provided positive evidence of diabetes or diabetes treatment. A negative label was assigned when `DIQ010`=2 and neither treatment indicator was positive. Borderline `DIQ010` responses were excluded unless `DIQ050` or `DIQ070` provided positive evidence.

The full fasting cohort was divided into training, validation, and test sets. Model fitting and rule generation used training data, model and rule selection used validation data, and the test set was reserved for final evaluation. The selected binary rules were subsequently weighted with a nonnegative survey-weighted logistic model. For participant *i*, the DMN score was

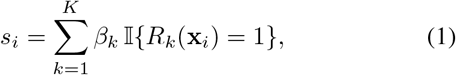

where *β*_*k*_ ≥ 0 and I {·} denotes rule activation. The probability mapping was

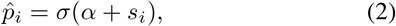

where *σ*(*z*) = 1/(1 + exp(− *z*)).

For counterfactual analysis, BMI was reduced while all other variables were held fixed. The DMN was re-executed and the probability difference was computed using the frozen mapping in Eq. (2). The analysis therefore quantifies sensitivity of the deployed decision policy and not the causal effect of BMI reduction.

## IV. Experimental Evaluation

### A. Data preprocessing

An analysis-ready dataset was constructed from NHANES Cycle J by merging demographic, examination, laboratory, and questionnaire modules. Repeated blood-pressure measurements were summarized by their row-wise means. Smoking responses were grouped into interpretable categories. Missing predictor values were imputed using training-set medians within each fitted pipeline. Clinically meaningful biomarkers were retained without outlier clipping.

The full fasting analysis cohort contained 2,582 participants: 472 positive and 2,110 negative cases. Its survey-weighted label prevalence was 12.43%. Gate0 was defined as HbA_1*c*_ < 6.5% and fasting plasma glucose (FPG) < 126 mg*/*dL while retaining both positive and negative reference labels. Gate0 contained 2,111 participants: 119 positive and 1,992 negative cases, with a survey-weighted prevalence of 3.15%.

The analysis cohort was split 60/20/20, stratifying jointly by the outcome and Gate0 membership. The training, validation, and test sets contained 1,549, 516, and 517 participants, respectively. Their Gate0 subsets contained 1,266, 422, and 423 participants, including 71, 24, and 24 positive cases. The development cohort, defined as training plus validation, therefore contained 2,065 participants, including 1,688 in Gate0. Participant identifiers were mutually exclusive across the three partitions.

### B. Feature screening

A survey-weighted L2-regularized logistic model was fitted on the training set. The regularization parameter was selected by five-fold stratified cross-validation within the training partition using weighted log loss. The evaluated grid was *C* ∈ {0.01, 0.03, 0.05, 0.08, 0.1, 0.2, 0.5, 1, 2, 5, 10}; the selected value was *C* = 0.01. After refitting on the complete training set, predictor relevance was assessed by the mean decrease in weighted validation ROC–AUC over 100 independent permutations per feature [16]. All random operations used seed 42. As a complementary, unweighted screening measure, mutual information was computed on the training set. Permutation importance measures the conditional contribution of a feature to the fitted model, whereas mutual information measures marginal dependence with the outcome.

HbA_1*c*_ was dominant under both measures. Total cholesterol and triglycerides made the largest additional conditional contributions in the fitted reference model. FPG had high mutual information but near-zero incremental permutation importance, consistent with strong overlap with HbA_1*c*_. Age, anthropometric variables, blood pressure, and lipid markers were retained as candidate families for interpretable rule construction. Small negative permutation estimates were treated as sampling variation around zero rather than evidence of protective effects.

### C. Rule-ensemble model

A RuleFit-style rule ensemble was fitted using the training set [17]. The implementation generated 400 shallow gradient-boosted trees with learning rate 0.03, a maximum of four leaves per tree, minimum leaf size 10, subsampling rate 0.70, and random seed 42. Terminal-node paths were extracted as binary rules and combined with standardized linear terms in an L1-regularized weighted logistic model. Candidate values *C* ∈ {0.03, 0.05, 0.08, 0.10, 0.20, 0.50, 1.00} were compared using weighted validation log loss; the selected value was *C* = 0.50. The fixed configuration was then refitted on the combined training and validation data and evaluated once on the untouched test set. Fasting-subsample weights were used directly during tree fitting, logistic fitting, and metric calculation.

### D. Clinical candidate-rule selection

A clinically constrained library of 172 three-condition rules was constructed from a prespecified threshold set supported by the training analyses. The library comprised HbA_1*c*_ 5.7– 6.4% and 6.0–6.4%; FPG 100–125 and 110–125 mg/dL; BMI ≥ 25 and ≥ 30 kg/m^2^; blood pressure ≥ 130/80 mmHg; triglycerides ≥ 150 mg/dL; sex-specific low HDL; total cholesterol ≥ 200 mg/dL; LDL ≥ 130 mg/dL; and age ≥ 65 and ≥ 75 years. Every candidate contained three distinct variable families and at least one glycemic condition; combinations containing two thresholds from the same family were excluded. This clinicalization stage was separate from the rule-ensemble fit, and the thresholds were not presented as literal reproductions of individual tree cut-points.

Selection was performed exclusively in the Gate0 validation subset. A candidate was eligible when its survey-weighted support was at least 2%, it covered at least 15 participants, included at least three positive cases, and had lift greater than one. Within each distinct feature-family profile, the rule with the highest validation confidence was retained. Four profiles were frozen before evaluation on the test set. Support, confidence, and lift were defined as

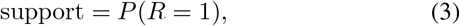

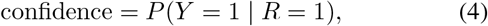

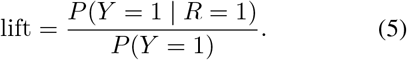

Table III summarizes the untouched-test statistics, while Panel A of Table IV preserves the corresponding rule conditions. The wide intervals, particularly for C2 and C4, reflect the limited number of positive test cases. These rules should therefore be interpreted as risk-enrichment signals requiring clinical and external validation rather than as definitive risk strata.

**TABLE I.** Feature screening on the training and validation partitions.

| Feature | Mean AUC drop | SD | MI |
| --- | --- | --- | --- |
| HbA<sub>1c</sub> | 0.2137 | 0.0367 | 0.2344 |
| Total cholesterol | 0.0634 | 0.0191 | 0.0057 |
| Triglycerides | 0.0274 | 0.0077 | 0.0294 |
| LDL cholesterol | 0.0115 | 0.0065 | 0.0169 |
| Age | 0.0058 | 0.0047 | 0.0714 |
| Systolic BP | 0.0025 | 0.0019 | 0.0013 |
| Waist circumference | 0.0011 | 0.0030 | 0.0289 |
| BMI | 0.0009 | 0.0024 | 0.0217 |
| Smoking status | 0.0008 | 0.0015 | 0.0076 |
| Diastolic BP | 0.0005 | 0.0036 | 0.0000 |
| hs-CRP | 0.0003 | 0.0014 | 0.0000 |
| FPG | −0.0006 | 0.0024 | 0.1914 |
| Sex | −0.0008 | 0.0013 | 0.0003 |
| HDL cholesterol | −0.0009 | 0.0039 | 0.0169 |
| Income-to-poverty ratio | −0.0034 | 0.0016 | 0.0174 |

**TABLE II.**
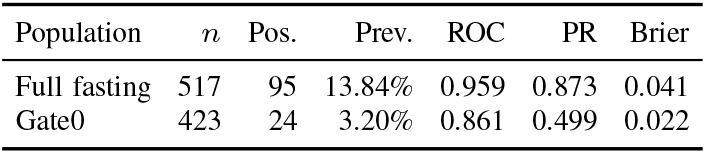
Untouched-test performance of the rule-ensemble model.

| Population | $n$ | Pos. | Prev. | ROC | PR | Brier |
| --- | --- | --- | --- | --- | --- | --- |
| Full fasting | 517 | 95 | 13.84% | 0.959 | 0.873 | 0.041 |
| Gate0 | 423 | 24 | 3.20% | 0.861 | 0.499 | 0.022 |

**TABLE III.** Untouched Gate0 test statistics for the four validation-selected candidate rules.

| ID | $n$ | Pos. | Support | Lift | Confidence, % (95% CI) |
| --- | --- | --- | --- | --- | --- |
| C1 | 23 | 7 | 2.80% | 8.85 | 28.3 (6.4–53.1) |
| C2 | 10 | 4 | 0.88% | 11.25 | 36.0 (0.0–48.5) |
| C3 | 15 | 5 | 1.13% | 10.38 | 33.2 (4.7–61.0) |
| C4 | 29 | 2 | 5.68% | 6.22 | 19.9 (0.0–36.5) |

Note: Support, confidence, and lift are survey-weighted; $n$ and Pos. are unweighted counts. The complete rule conditions are shown in Panel A of Table IV.

**TABLE IV.**
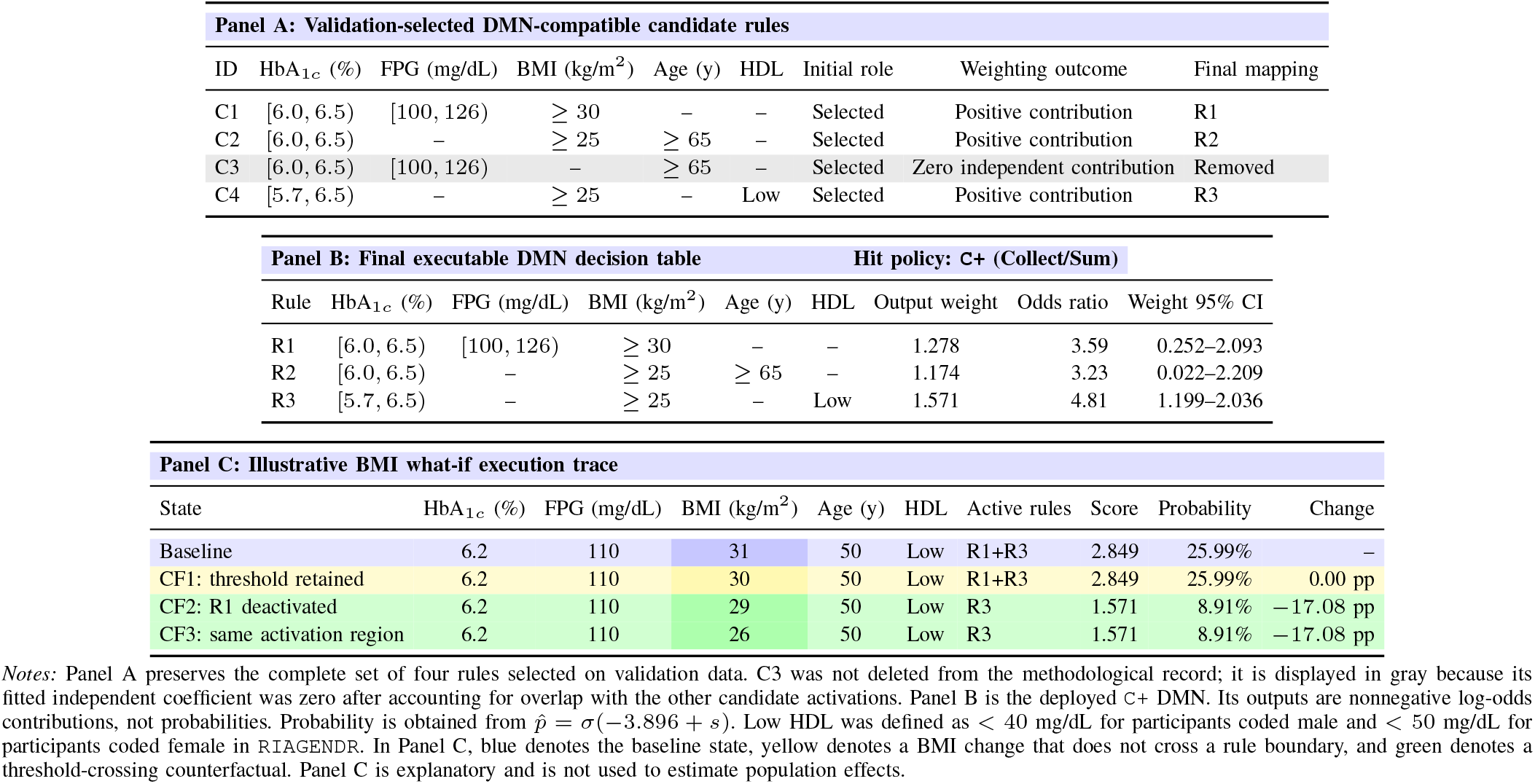
Traceable DMN representation and BMI what-if execution.

### E. Final DMN representation and probability mapping

The four validation-selected candidate activations shown in Panel A of Table IV were fitted in the combined training-and-validation Gate0 cohort using a survey-weighted logistic model with nonnegative coefficients. The nonnegative constraint preserved the collect/sum interpretation: activation of a risk-enrichment rule could not lower the score. Candidate C3 received a zero independent coefficient after accounting for overlap with the other activations and was removed. The final DMN therefore contained three rules.

Table IV provides the central visual representation of the framework. Panel A preserves all four validation-selected candidate rules in DMN-compatible form. Candidate C3 is retained for traceability but shaded in gray because its independent coefficient was estimated as zero in the nonnegative weighting model. Panel B shows the resulting executable DMN with the `C+` hit policy (Collect with Sum aggregation). Panel C provides a color-coded BMI what-if execution trace.

The final score and probability mapping were

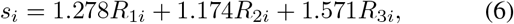

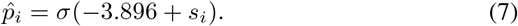

When none of the rules fired, the estimated probability was 1.99%. When all three fired, the fitted probability was 53.18%. These values are outputs of the fitted mapping and not observed subgroup prevalence estimates.

On the untouched Gate0 test set, the final DMN achieved ROC–AUC 0.769, PR–AUC 0.153, Brier score 0.029, and weighted log loss 0.122. An intercept-only comparator had ROC–AUC 0.500, PR–AUC 0.032, Brier score 0.031, and log loss 0.142. The simpler DMN was less discriminative than the full rule ensemble, as expected, but provided a compact and auditable policy representation.

### F. Counterfactual BMI sensitivity

BMI was selected because it appeared in all three final rules and could change the DMN through thresholds at 30 and 25 kg/m^2^. In the untouched Gate0 test subset, BMI was reduced by 1, 3, or 5 kg/m^2^, with values bounded below by 18.5 kg/m^2^. All other variables were held fixed. The frozen DMN was then re-executed and the same probability mapping was applied.

Because the DMN uses binary threshold conditions, the response was stepwise. A BMI reduction produced no probability change unless it deactivated at least one active rule. Mean changes were calculated within subgroups in which each rule fired at baseline. Uncertainty was estimated using 1,000 bootstrap replicates that resampled NHANES primary sampling units within strata while retaining the original fasting-subsample weights.

An isotonic mapping fitted on development scores was used as a robustness check. Both mappings showed the same directional pattern: larger reductions crossed more BMI thresholds and deactivated more rules, although absolute probability estimates differed because the score had only a small number of discrete activation patterns (Fig. 3).

**Fig. 3.**
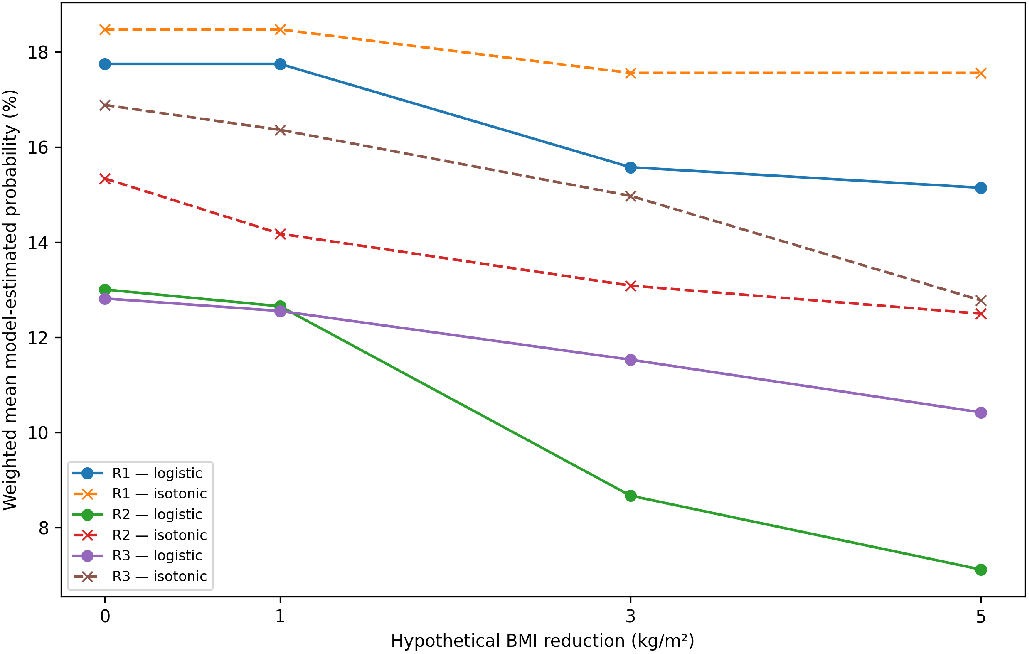
Survey-weighted logistic and isotonic probability mappings under hypothetical BMI reductions.

Panel C of Table IV visualizes the execution mechanism. At BMI 31 and 30 kg/m^2^, both R1 and R3 remain active, so the score and probability are unchanged. Reducing BMI below 30 kg/m^2^ deactivates R1 while R3 remains active, producing a discrete probability change from 25.99% to 8.91%. Further reductions within the interval 25 ≤ BMI < 30 do not change the active-rule profile. This colored trace is explanatory and is not used to estimate the population-level effects reported in Table V.

**TABLE V.**
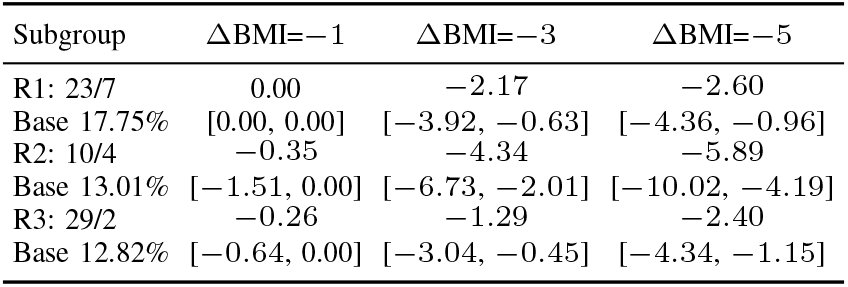
Survey-weighted BMI what-if results in Gate0 test subgroups.

| Subgroup | $\Delta\text{BMI}=-1$ | $\Delta\text{BMI}=-3$ | $\Delta\text{BMI}=-5$ |
| --- | --- | --- | --- |
| R1: 23/7 | 0.00 | –2.17 | –2.60 |
| Base 17.75% | [0.00, 0.00] | [–3.92, –0.63] | [–4.36, –0.96] |
| R2: 10/4 | –0.35 | –4.34 | –5.89 |
| Base 13.01% | [–1.51, 0.00] | [–6.73, –2.01] | [–10.02, –4.19] |
| R3: 29/2 | –0.26 | –1.29 | –2.40 |
| Base 12.82% | [–0.64, 0.00] | [–3.04, –0.45] | [–4.34, –1.15] |

### G. Reproducibility

All data partitions were fixed before model fitting, participant identifiers were mutually exclusive across partitions, and random operations used seed 42 unless otherwise stated.

## V. Discussion

The results illustrate the distinction between a predictive rule-ensemble model and an executable DMN policy. The full model retained many linear and rule terms and achieved higher test discrimination. The DMN deliberately sacrificed predictive detail to obtain three inspectable rules, additive nonnegative contributions, and a finite set of possible activation profiles.

The counterfactual analysis is most naturally interpreted as policy sensitivity. It identifies the threshold crossing responsible for a changed output and makes clear when no change occurs. This is preferable to reporting smooth probability reductions that are inconsistent with binary DMN rule activation. The analysis can therefore support transparent review of how a deployed decision table behaves, but it does not establish that reducing BMI would cause the estimated change in diabetes status or future disease risk.

The candidate-rule confidence intervals were wide because Gate0 contained only 24 positive cases in the untouched test subset. The reported intervals condition on the selected rule set and therefore do not represent the additional uncertainty introduced by screening 172 candidate rules. The reported enrichments and probability mapping should be validated in larger and independent datasets before clinical use.

## VI. Limitations and Future Work

This study has several limitations. First, the analysis used only one NHANES cycle, resulting in a small number of positive cases, particularly in the Gate0 test set. This limited the precision and generalizability of the rule estimates. Second, the diabetes reference label partly relied on self-reported questionnaire and treatment information rather than laboratory measurements alone, which may introduce reporting or outcome-misclassification bias. In addition, because NHANES is cross-sectional, the identified rules describe associations with documented diabetes status and do not predict future diabetes incidence.

The candidate rules were clinically constrained but were not derived from formal guideline encoding or reviewed by a clinical expert panel. The confidence intervals also condition on the selected rule set and do not include all uncertainty introduced during rule generation and selection.

Future work will combine multiple NHANES cycles to increase the number of positive cases and will evaluate the framework on independent datasets and different populations. Clinical expert review and prospective evaluation will also be needed before the rules can be used as an early-alert mechanism in practice.

## VII. Conclusion

This paper presented a validation-separated framework that combines survey-weighted rule-ensemble learning, executable DMN logic, and counterfactual policy-sensitivity analysis. In the NHANES case study, the predictive rule ensemble identified combinations of clinical risk factors associated with documented diabetes status, while the final three-rule DMN provided a compact, transparent, and auditable representation of this evidence. The BMI what-if analysis further showed that DMN outputs changed only when a hypothetical reduction crossed an explicit rule threshold, making the mechanism behind each probability change traceable.

The derived rules are not proposed as new clinical guideline recommendations or diagnostic criteria. Instead, they demonstrate how data-supported DMN logic could be used as an early-alert mechanism for individuals who remain below diagnostic glycemic thresholds but present combinations of risk factors associated with a higher estimated probability of documented diabetes status. Such an alert could support closer monitoring, additional testing, or earlier clinical review, while leaving diagnosis and treatment decisions to qualified healthcare professionals.

Overall, the framework extends static decision logic with patient-specific and interpretable what-if reasoning while maintaining a clear distinction between predictive association, policy sensitivity, and causal effect. External validation, clinical expert review, and prospective workflow evaluation remain necessary before the framework can be considered for clinical deployment.

## Data Availability

The de-identified National Health and Nutrition Examination Survey (NHANES) data analyzed in this study are publicly available from the National Center for Health Statistics. The study used publicly released demographic, examination, laboratory, and questionnaire data. No restricted-access data were used.

https://wwwn.cdc.gov/nchs/nhanes/continuousnhanes/default.aspx?BeginYear=2017

## Ethics Approval and Consent to Participate

This study used publicly available, de-identified data from the National Health and Nutrition Examination Survey (NHANES). NHANES protocols were approved by the National Center for Health Statistics Research Ethics Review Board, and informed consent was obtained from participants. No new participant recruitment or data collection was conducted.

## Author Approval

All authors reviewed and approved the manuscript and consented to its submission and posting as a preprint on medRxiv.

## Data Availability

The de-identified NHANES data analyzed in this study are publicly available from the National Center for Health Statistics.

